# Spatial Transcriptomics Recontextualizes the Cellular Environment of Conjunctival Melanoma

**DOI:** 10.64898/2026.06.23.26356337

**Authors:** Jeffrey Maurer, Yoko Suzuki-Horiuchi, Bryant Duong, Michael Vedi Ramirez, Albert Chen, Stephen M. Prouty, Tatyana Milman, Vivian Lee, Yuyan Cheng

**Author notes:** Corresponding authors, Corresponding Author: Yuyan Cheng, E-mail address; Co-Corresponding Author: Vivian Lee. These authors contributed equally (Jeffrey Maurer, Yoko Suzuki-Horiuchi).

## Abstract

**Introduction:** Conjunctival melanoma (CM) is a rare cancer with a potentially high recurrence rate. The mechanics of its progression, its relationship with neighboring tissues, and its molecular characteristics are largely unknown. Diagnosis currently requires a biopsy and the time and expertise of a pathologist.

**Methods:** Archived human biopsies containing CM were submitted to Xenium spatial transcriptomic analysis. Regions were graded by disease progression through histopathology. Differential expression (DE) and composition analysis were performed across disease states.

**Results:** From three patients, 12 formalin-fixed paraffin-embedded (FFPE) tissue specimens were recovered. Composition analysis showed that melanoma depletes fibroblast and epithelial cells while melanocytes proliferate. DE signatures specific to each state show a clear pattern of progression from inflammation, to cellular restructuring, and then to tumor progression and malignancy.

**Conclusion:** Spatial transcriptomics allows single-cell transcriptomics techniques to compare spatially relevant annotations that are difficult to separate by library. This study proposes disease progression biomarker candidates that may elucidate the mechanics of CM progression and function as objective diagnostic and prognostic tools in the future.

**Plain Language Abstract:** Conjunctival melanoma is a rare cancer of the eye. Ideally, its diagnosis, prognosis, and treatment would be guided by measurable biomarkers. However, definitive markers are lacking, and no ancillary studies, such as immunohistochemical stains or molecular studies like bulk RNA-seq, can reliably distinguish benign from premalignant/malignant lesions, predict outcomes, or guide treatment. As a result, diagnosis frequently relies heavily on morphologic assessment by an experienced pathologist. To address this gap, biopsied tissues from three patients with conjunctival melanoma were obtained for this study. Regions of the tissue were manually labeled from normal, healthy tissue through three stages of increasing cancer severity. With thin tissue slices, new spatial transcriptomic technology measured the expression of genes across the tissue. Previous methods would have required the annotated tissue to be carefully physically separated before measuring expression. Comparing cells labeled as healthy to melanoma, there is a sharp change in the proportion of cell types present. The cell types that are present in the tumor also express genes differently compared to their counterparts outside of the tumor. Some of these genes only change during one portion of tumor progression. Measuring the levels of these severity-dependent genes may provide an alternative diagnostic method. Given the role of these genes, it is also possible to infer how conjunctival melanoma develops and becomes malignant to inform the development of future treatments.

## Introduction

Conjunctival melanomas are malignant tumors arising from conjunctival melanocytes. Tumors develop either de novo, or from benign melanocytic nevi or premalignant lesions, referred previously as primary acquired melanosis (PAM) and currently as conjunctival melanocytic intraepithelial lesions (CMIL) (1). Although conjunctival melanomas account for only 5% of ocular melanomas (2) and 0.25% of all melanomas (3), their incidence has been rising, with reports of doubling in certain populations over the past two decades (4–6). Despite treatment, these tumors carry substantial morbidity and mortality, with 10-year estimates of vision loss approaching 45%, local recurrence or new tumor formation at 64%, and disease-related death at 33% (7). Notably, when matched for size, conjunctival melanomas carry a higher mortality rate than uveal melanomas, underscoring the gravity of this disease (8).

Although histopathology remains the most reliable diagnostic method, conjunctival melanoma can be difficult to diagnose, particularly when lesions display ambiguous or atypical histologic features (1). Similarly, the potential of such lesions to recur or metastasize following complete excision remains difficult to predict on morphologic grounds alone. Unlike uveal melanomas and retinoblastomas, for which molecular biomarkers now guide risk stratification and therapeutic selection, few biomarkers currently exist for conjunctival melanocytic lesions (9) (10). Extrapolation from other melanocytic tumors is of limited utility, as conjunctival melanomas have a distinct biology. Even shared mutational signatures with cutaneous melanomas are mostly restricted to general UV-damage markers (9, 11, 12).

Transcriptional profiling has proven valuable in disclosing the molecular mechanisms underlying tumorigenesis in a wide range of malignancies. In particular, spatial transcriptomic methods that preserve the tissue architecture are critical for understanding tumor heterogeneity. The tumor microenvironment harbors clonal populations residing in specialized niches with differentially enriched transcriptional programs that regulate metabolism, inflammation, motility, and proliferation—features that influence invasion, metastasis, and therapeutic response. Remarkably, the transcriptional landscape of conjunctival melanocytic lesions has remained largely uncharacterized, owing principally to the disease rarity, limited tissue availability, and compact tumor size with the latter two precluding conventional RNA sequencing approaches (13).

To address this gap, we employed 10X Genomics Xenium, a platform that generates high-plex in situ data from spatially defined regions of interest within formalin-fixed, paraffin-embedded tissue. We report the first spatial transcriptomic characterization of conjunctival melanomas arising from CMIL, identifying differentially expressed transcriptional pathways that illuminate the molecular mechanisms mediating conjunctival melanoma tumorigenesis and provide novel candidate biomarkers for this understudied disease.

## Methods

### Human Materials

#### Sample Acquisition

Twelve formalin-fixed paraffin-embedded (FFPE) tissue sections containing conjunctival melanoma (CM) and adjacent conjunctival melanocytic intraepithelial lesion (CMIL) and/or unremarkable conjunctiva were retrieved from the pathology archives at the Wills Eye Hospital, Philadelphia (IRB protocol #2023-64). Each case was recut and stained with hematoxylin and eosin (H&E) to reconfirm the diagnosis of CM and/or CMIL. Tissue biopsies were re-embedded in a single composite block for spatial transcriptomics (Figure 1). The composite block was cut under RNase-free conditions.

**Fig. 1.**
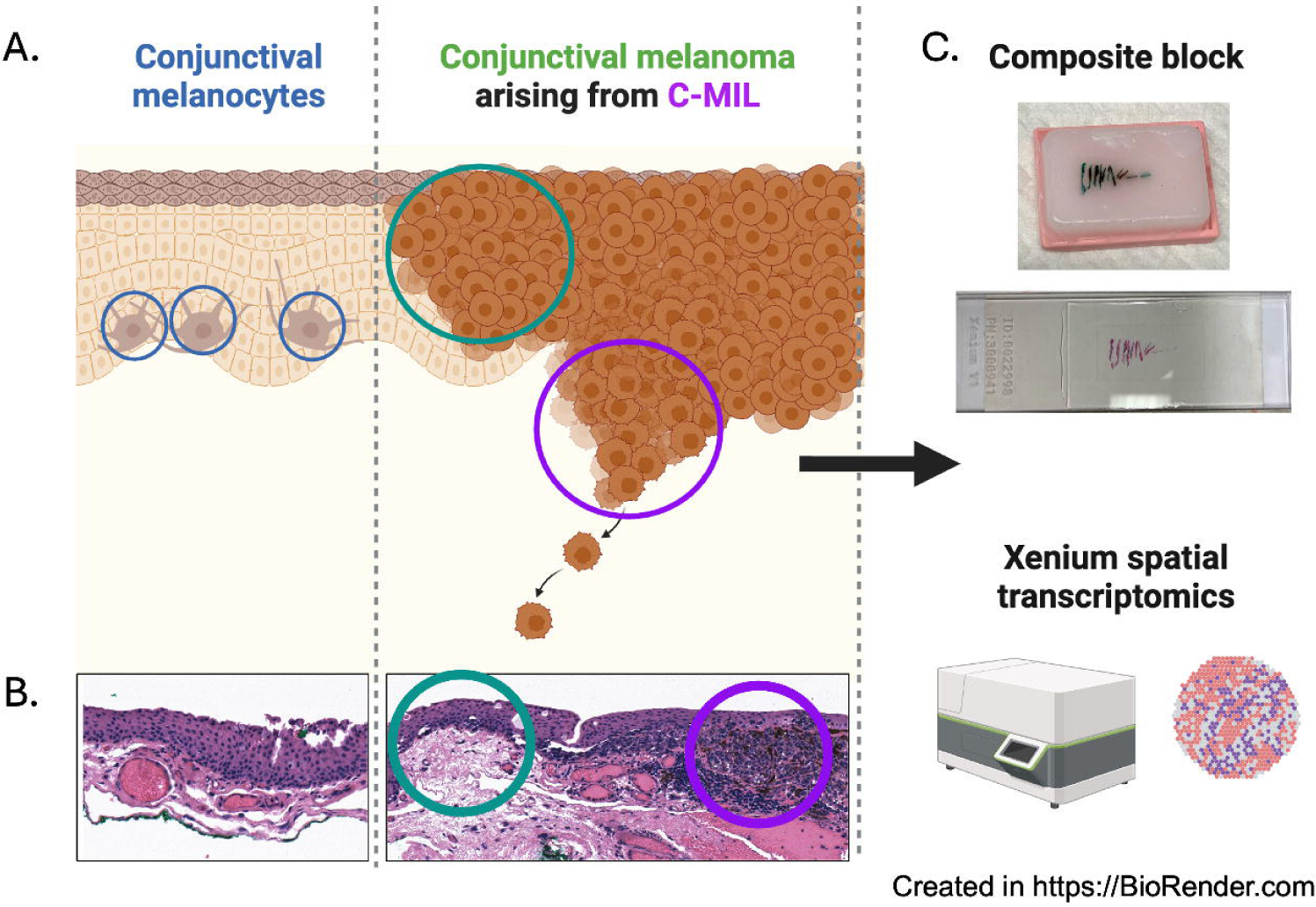
Schematic of study design. (a) Diagram illustrating unmarkable conjunctival melanocytes (blue circles), CMIL (green circle), and melanoma (purple circle). (b) Photomicrograph of representative H&E sections correlating with disease subtypes – normal, CMIL (green circle), and melanoma (purple circle). (c) Study design with images of composite block and slide.

#### Tissue Composite Block and Staining

For confirmation of diagnosis and case selection, slides were stained using standard H&E procedures. For placement on the Xenium slide (10X Genomics, Pleasanton CA), the individual tissues were melted out of their FFPE blocks and assembled into a single composite FFPE block with the tissues aligned to ensure that all samples would fit within the Xenium sample area. Using nuclease-free microtomy, a ribbon of 5-micron sections was cut from the composite block and floated on a water bath at 43°C. A single section was selected and attached to the Xenium slide, ensuring that the tissues did not overlap the fiducials. The Xenium slide was dried at room temperature for 1 hour, then for 3 hours at 42 degrees Celsius, before storage in a desiccator at room temperature.

#### Imaging

Imaging was performed using Keyence (BZ-X700) and Leica Aperio Versa Slide Scanner. Stitched images were taken at 20X magnification.

#### Xenium in situ Spatial Transcriptomics

For Xenium in situ spatial transcriptomics, the 10X Genomics protocol was used (Protocol #: CG000580, CG000582, CG000584). Briefly, slides were baked for 2 hours at 60°C on thermal cycler using a Xenium v1 thermocycler adaptor (PN-3000954) followed by deparaffinization in xylene and rehydration with ethanol gradient. Slides were de-crosslinked at 80°C for 30 min, then at 22°C for 10 min. mRNAs were targeted with the Xenium Human Skin Gene Expression Panel (PN-100643). Probes were preheated by incubating for 2 min at 95°C then placed on ice. Hybridization was performed overnight (16 - 24h) at 50°C in thermal cycler. The following day, post-hybridization wash was performed at 37°C for 30 min. Probes were ligated at 37°C for 2 hours and amplified at 30°C for 2 hours with PBST washes in between. Slides underwent staining with reducing agent, autofluorescence quenching, and ethanol washes, then dried at 37°C for 5 min. Nuclei were stained with DAPI for 1 min. Xenium imaging processing, decoding, quality score generation, and DAPI-based segmentation were performed using the Xenium Analyzer.

#### H&E Staining

After the Xenium run, H&E staining was performed on the Xenium slide. The slide was removed from the cassette containing phosphate-buffered saline-tween 20 (0.05%) and placed in a Coplin jar containing 50 ml of 10 mM sodium hydrosulfite (Sigma, 157953-1KG) solution and incubated for 10 minutes at room temperature. The slide underwent multiple rounds of washing in between incubation with hematoxylin (Gill 3, Surgipath, 3801540), bluing solution (Azer Scientific, ES745-1G), and eosin (Surgipath, 3801600). The slide was then dehydrated in 95% (Azer Scientific, ES753) and 100% (Azer Scientific, ES631) ethanol, cleared in xylene substitute (Azer Scientific, ES657), then coverslipped using MM24 mounting medium (Surgipath, 3801120).

#### H&E Image Alignment and Annotation in Xenium explorer

H&E sections were imaged with a slide scanner, converted to ome.tiff file using QuPath v0.5.1, and aligned using the 10x Genomics Xenium Explorer (14). The image was aligned to the DAPI staining image in Xenium Explorer. Manual annotation was first performed by a pathologist on H&E image. Then, regions of interest were selected in Xenium Explorer.

### Segmentation Analysis

#### Default 10X

The 10x Genomics Xenium Analyzer used the 10x Genomics Xenium Onboard Analysis v1.7 - v1.9, major version 3.0 (14) to call transcripts, assign Q-scores, and perform a naïve segmentation algorithm. This algorithm expands DAPI nucleus calls by fifteen microns.

#### Cellpose

Cellpose (15) used the highest resolution of morphology_mip.ome.tif DAPI stain to perform segmentation. Cellpose was run using the SOPA v2.1.11 API (16). The parameters used are reported in the Github repository but are stated here briefly. The image was divided into square patches of 1024 pixels with 256 pixels of overlap. The model_type was set to ‘nuclei’ and diameter set to 35 pixels for the expected diameter of cells.

#### Baysor

Control probes and transcripts with a Q-score below 20 were filtered out. Baysor v0.6.2 (17) performed segmentation with JULIA_NUM_THREADS=8 (18) and the following parameters that are also reported in the config file with parameters on the Github repository under ‘config’. In the data section, min_molecules_per_gene set to 1 and min_molecules_per_cell set to 12. In the segmentation section, n_clusters set to 10, prior_segmentation_confidence set to 0.9, iters set to 500 and the n_cells_init set to the estimate by 10X genomics estimate using DAPI which was 108615. Lastly, the plotting variable min_pixels_per_cell was set to 5.

The 10x Genomics Xenium Ranger v1.7.1.1 then assigned transcripts to the segmented cells and formatted the output to match the original output format.

#### Mutually Exclusive Co-expression Rates (MECR)

To compare the segmentations, the MECR (19) of canonical cell markers were calculated with formula [1].

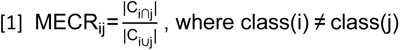

This is where *i* and *j* are a pair of genes that are markers for different cell classes, |C_i∩j_| in the numerator is the number of cells in our dataset that express both markers and |C_iUj_| in the denominator is the number of cells that express at least one of the two markers. The MECR distribution of each segmentation dataset was compared with a Wilcoxon signed-rank test.

### Cell Processing

The raw count matrix from Baysor segmentation was processed using the R v4.3.2 Seurat package v5.2 (20). Raw counts were normalized using SCTransform(). The top 50 principal components were calculated from all 260 target genes with RunPCA(). This was followed by FindNeighbors() and RunUMAP() where k.param and n.neighbors parameters were set to 50 for their respective function.

### Tissue and Cell Annotation

#### Clustering and Cell Type Annotation

Clustering with FindClusters() used the Leiden algorithm at a variety of resolutions (0.05, 0.10, 0.25, 0.35, 0.5). As a heuristic, a resolution with more clusters than the expected number of biological groups is chosen. Expecting around seven biological cell groups this sample, a resolution of 0.35 with 12 clusters was chosen to annotate. Canonical cell markers are listed here: B Cell: CD79A, MZB1, TNFRSF17; T Cell: CD3G, IL32, LCK; Macrophage: CD68, C5AR1; Melanocyte: MLANA, PMEL, BCAN; Epithelium: AHNAK2, DSC1, IGFBP3, KRTDAP, LY6D, LYPD3, PCDH7, SERPINB5; Fibroblast: COL5A2, COL6A1, COL6A2. Using expression of these markers and the results of Seurat’s FindAllMarkers(), clusters were labelled as defined cell types. There were four populations of unclear identity comprising ∼15.6k cells. Given the limited collection of only 260 genes, cell typing these populations without additional markers is difficult. Further analysis omits these cells unless otherwise specified.

#### Disease State Annotation

Spatial disease annotation was performed using the selection tool within Xenium Explorer v3-v4 (14). The bounded regions of disease assignment were overlayed onto cells using the sp v2.2-0 package in R (21). This was repeated for patient annotation. Future analysis uses the 110,026 cells with a complete annotation of disease stage, patient of origin and cell type.

### Cell Composition Analysis

Cell composition analysis was performed by scCODA v0.1.9 with python v3.11.7 (22). Composition was split by disease, cell type and patient of origin. The model fit disease using the healthy annotated cells as the reference level. The total cells per patient was used as a reference “cell type” required by the tool. A pseudocount of 0.5 was added to all values. The Hamiltonian Monte Carlo sampling used 80k cycles with 20k of burn-in, resulting in a sampling acceptance rate of 38.2%. The default desired False Discovery Rate (FDR) of 0.05 and 94% High Density Interval (HDI) were used to call significant changes in composition. The scCODA package does not calculate p-values.

### Differential Gene Expression Analysis

Differential expression analysis between each disease state from all others was performed on pseudobulk samples generated with Seurat’s AggregateExpression() across patient, cell type and disease state. These pseudobulk samples were assessed using DESeq2 v1.42.0 (23), limma v3.58.1 (24) and EdgeR v4.0.14 (25). Each cell type was assessed separately but with the same model formula: ∼0 + disease. All contrasts were calculated for each cell type using each method. Multiple testing correction was omitted for this analysis due to limited sample size. A differentially expressed gene (DEG) was defined as a gene that any of the three methods called as significant using a raw p-value threshold of 0.05 with no effect size threshold.

## Results

### Clinical and Histopathologic Characteristics

Twelve tissue samples were collected from three patients with conjunctival melanoma. Patient demographics are presented in Supplementary table 1. Tissue biopsies included the regions diagnosed with CM, CMIL, as well as healthy tissue (Figure 1).

### Baysor Optimally Segments Cells

Accurate cell segmentation is critical in spatial transcriptomics analysis because all downstream analyses—including cell typing, clustering, and spatial region annotation—depend on correctly defining cell boundaries and assigning transcripts. To evaluate segmentation performance in our dataset, we compared the default 10X segmentation with two alternative approaches, Cellpose (15) and Baysor (17) (Fig. 2. a). Cellpose was included as a widely used image-based method, whereas Baysor was selected because it uses a different algorithm that relies on transcript spatial information for segmentation. Visual inspection showed that both Cellpose and Baysor more clearly delineated intercellular boundaries and reduced assignment of noise in extracellular space to nearby cells compared to the default 10X segmentation (shown by arrows in Fig. 2. b).

**Fig. 2.**
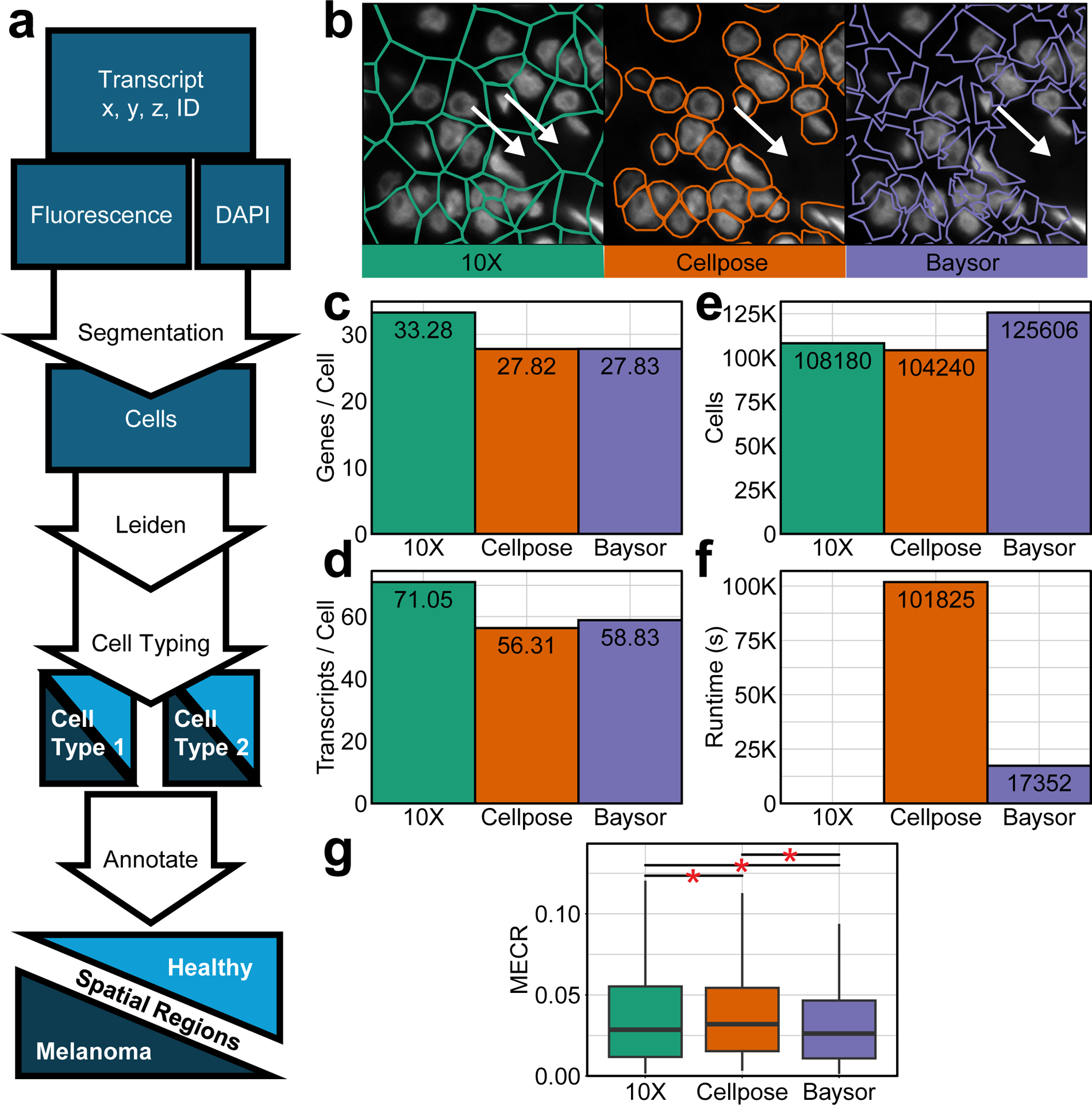
Baysor improves segmentation specificity and cell recovery for conjunctival melanoma spatial transcriptomics. (a) Schematic of the spatial transcriptomic analysis workflow, from transcript coordinates and imaging inputs through segmentation, clustering, cell typing and spatial region annotation. (b) A representative 50x50 micron region showing 10X, Cellpose and Baysor’s segmentation algorithms defining called cells’ boundaries overlayed on the DAPI signal. The arrows indicate a region of disagreement between 10X and the other algorithms. (C) Comparison of genes per cell across segmentation methods. (d) Transcripts per cell across segmentation methods. (e) Total number of cells identified by each method. (f) Runtime comparison of each method. (g) Distribution of MECR scores for cells segmented by each method using 22 cell markers of six cell types (B Cell, T Cell, Melanocyte, Fibroblast, Lymphocyte, Epithelium).

We next compared segmentation algorithm performance using genes per cell, transcripts per cell, total cells identified, runtime, and marker exclusivity. The default 10X pipeline yielded the highest mean genes per cell, 33.28, and transcripts per cell, 71.05, compared with Cellpose—27.81 and 56.31 respectively—and Baysor—27.83 and 58.83 respectively (Fig. 2. c-d). This likely reflects the loss of extracellular transcripts assigned to cells. Baysor identified the most cells, followed by 10X and then Cellpose (Fig. 2.e). Baysor was also much faster than Cellpose, with a runtime that allows same-day results (Fig. 2. f).

To directly assess segmentation specificity, we quantified the mutually exclusive co-expression rate (MECR), which measures the extent to which canonical markers from distinct cell types are incorrectly detected within the same segmented cell (Methods). Lower MECR indicates greater segmentation specificity and less transcript mixing across neighboring cells (19). Baysor had significantly lower MECR than 10X, indicating more specific transcript assignment (Fig. 2. g; Wilcoxon signed-rank test, p-value = 2.23e-32). Baysor also outperformed Cellpose (p-value = 5.01e-29), supporting its selection for downstream analyses based on overall specificity, cell recovery, and runtime.

### Cell Type Composition Changes with Disease State

Using Baysor-based segmentation, the dataset showed a median of 27 genes per cell, 45 transcripts per cell, and a cell area of 30 μm^2^ (Fig. 3. a). We performed downstream analysis on the segmented cells as described in Methods. Despite the relatively low depth, we resolved the major expected cell populations represented in the panel, including epithelial cells, melanocytes, fibroblasts, macrophages, T cells, and B cells (Fig. 3. b-c).

**Fig. 3.**
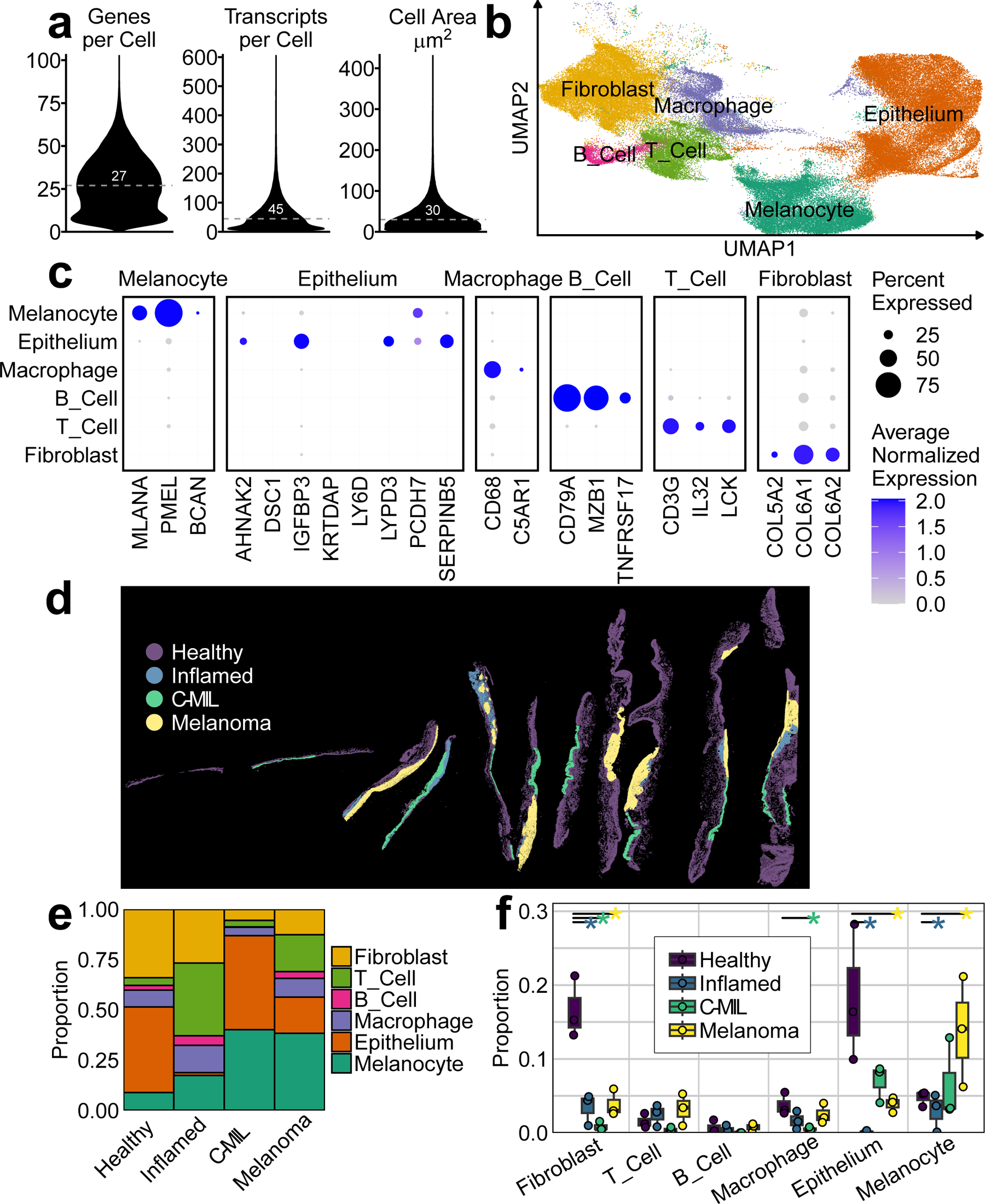
Cell-type annotation and disease-state-associated compositional changes in conjunctival melanoma. (a) Quality metrics after Baysor-based segmentation showing distributions of genes per cell, transcripts per cell and cell area. Median values are indicated. (b) UMAP projection of segmented cells showing the major annotated cell populations, including fibroblasts, T cells, B cells, macrophages, epithelial cells and melanocytes. (c) Dotplot showing the expression of canonical cell markers (d) Spatial map of manually annotated tissue regions classified as Healthy, Inflamed, CMIL and Melanoma. (e) Stacked bar plot showing relative cell-type composition across disease states. (f) A boxplot showing cell composition across disease state by patient. Bars above the boxes indicate significant differences compared to Healthy defined by scCODA.

Next, we manually annotated spatial regions into four disease states: healthy, inflamed, CMIL, and melanoma (Fig. 3. d). We then measured cellular composition across these regions. To test for significant compositional changes, we used scCODA, a Bayesian framework designed for differential cell composition even when sample sizes are limited (22). Compared with healthy regions, fibroblasts were reduced in inflamed (-2.118 to -0.747), CMIL (-3.092 to -1.391), and melanoma (-1.794 to -0.558) regions; epithelial cells were reduced in inflamed (-4.020 to -1.919), and melanoma (-1.722 to -0.439); and melanocytes were increased in melanoma (0.299 to 1.425) but reduced in inflamed (-1.897 to -0.364). Macrophages showed a weaker shift in CMIL (-1.645 to 0.013) (Fig. 3. e-f). These ranges represent the 94% credible interval for the estimated change relative to healthy tissue. Intervals entirely below zero indicate a decrease, intervals entirely above zero indicate an increase, and intervals spanning zero indicate uncertainty. Notably, melanoma regions contained significantly more melanocytes and fewer fibroblasts than healthy tissue, consistent with the expected cellular remodeling associated with melanoma progression. Together, this data resolves major cell classes and detects disease-state-associated shifts in tissue composition.

### Differential Gene Expression Changes Across Disease State

To identify stage-specific genes as candidate biomarkers, we performed pseudobulk differential expression (DE) analysis, aggregating cells by cell type, patient and disease state to reduce single-cell noise and increase robustness (Methods) (26). DE genes were called using a nominal p < 0.05 (Supplementary Table 2). Disease-specific DE gene sets for inflamed, CMIL, and melanoma were defined as genes significantly different in one pathological state compared to all other conditions (Methods). In the pooled analysis across all cell types, this yielded 39 inflamed-specific genes, 14 CMIL-specific genes, and 7 melanoma-specific genes (Supplementary Table 3).

These disease-specific gene sets highlight distinct transcriptional features across pathological states. Inflamed regions showed increased IL32 (Fig. 4. a), consistent with inflammatory signaling. CMIL was marked by reduced IGFBP7 in macrophages (Fig. 4. b). IGFBP7 is a secreted factor previously linked to senescence and tumor-suppressive responses in melanoma. Melanoma-specific changes included increased POSTN (Fig. 4. c), a matricellular gene associated with extracellular matrix remodeling, increased epithelial LY6D (Fig. 4. d), and decreased fibroblast MFAP5 (Fig. 4. e), indicating altered epithelial and stromal programs. Together, these stage-associated genes therefore nominate candidate spatial biomarkers for molecular discrimination of conjunctival melanoma progression.

**Fig. 4.**
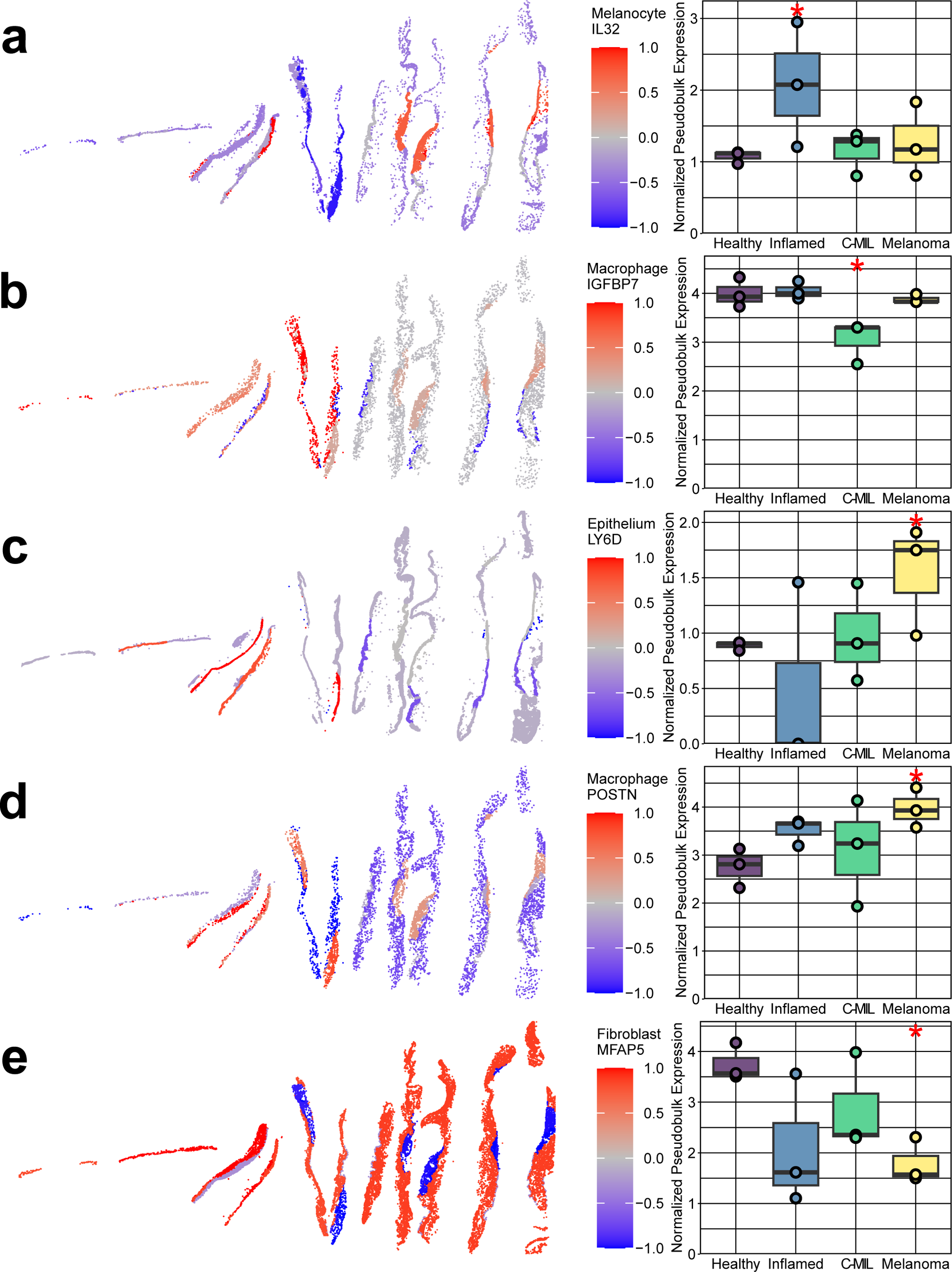
Stage-associated candidate spatial biomarkers across conjunctival melanoma disease states. Spatial expression maps and matched pseudobulk expression summaries for representative stage-associated genes identified by pseudobulk differential expression analysis. (a) IL32 in melanocytes. (b) IGFBP7 in macrophages. (c) POSTN in macrophages. (d) LY6D in epithelium. (e) MFAP5 in fibroblasts. For each panel, the left plot shows spatially resolved Z-normalized expression across tissue sections, and the right plot shows normalized pseudobulk expression across healthy, inflamed, CMIL and melanoma regions. A red asterisk above a condition’s box indicates that the gene’s expression for cells of that condition is significantly different from all other disease states (p < 0.05).

## Discussion

This study provides, to our knowledge, the first spatial transcriptomic view of conjunctival melanoma and its adjacent pathological states. Two landmark contributions emerge from this work, the generation of a new spatial transcriptomics dataset for a rare ocular malignancy and the establishment of a framework for interpreting conjunctival melanocytic lesions as spatially organized disease states with distinct molecular and cellular features. In this context, spatial transcriptomics offers a way to move beyond histopathology alone and towards a biologically grounded model of disease progression.

A central implication of this work is that conjunctival melanoma should be understood as a disease of tissue ecosystems rather than of melanocytes in isolation. The transitions across inflamed, CMIL and melanoma regions suggest progressive restructuring of the local epithelial, stromal and immune environment. Each state is defined by a different balance between inflammatory activation, tissue repair, stromal remodeling and malignant niche formation. This is conceptually important because it frames progression not simply as the accumulation of atypical melanocytes, but as stepwise reprogramming of the surrounding microenvironment into one that increasingly permits or supports tumor growth.

The molecular program identified here reinforces that reprogramming interpretation and suggests a stage-resolved biological model. The enrichment of IL32 in inflamed regions (Fig. 4. a) is consistent with a cytokine-driven activated tissue state, as IL32 is a pro-inflammatory cytokine that amplifies immune signaling and has been implicated broadly in inflammatory tissue responses (27). The CMIL state highlights a unique downregulation of IGFBP7 in macrophages (Fig. 4. b). Loss of IGFBP7 has been linked to melanoma progression and is widely regarded as a tumor-suppressive event in melanoma, particularly in the setting of oncogenic BRAF signaling (28, 29). In our data, reduced macrophage-associated IGFBP7 is therefore consistent with a transitional state in which inflammatory tissue begins to acquire features permissive for melanocytic progression. In melanoma, increased POSTN (Fig. 4. c) points to extracellular matrix remodeling within the tumor niche, a process increasingly recognized as a key determinant of melanoma progression and tumor-stroma crosstalk (30). Elevated epithelial LY6D (Fig. 4. d) further suggests acquisition of a more plastic epithelial state, as LY6D has been associated with squamous-like and therapy-resistant epithelial programs in skin tumors (31). By contrast, MFAP5 (Fig. 4. e) was downregulated in fibroblasts. Because MFAP5 is an extracellular matrix-associated fibroblast gene linked to activated stromal programs in multiple solid tumors (32, 33), its loss here may reflect the replacement of a normal structural fibroblast program by an alternative melanoma-associated remodeling state dominated by POSTN-rich stroma, rather than by uniform activation of all fibroblast matrix genes.

An important conceptual point is that cell abundance and cell state do not necessarily change in parallel. The melanoma-associated stromal program suggests that even when fibroblasts are not expanded, the fibroblasts that remain may be functionally reprogrammed in ways that are disproportionately important for tumor behavior. This distinction is likely to matter in CM, where local invasion, recurrence and interaction with the ocular surface depend heavily on tissue context (34). More broadly, it suggests that biomarkers of progression may be more informative when they capture spatial cell states and niche activity, rather than relying only on bulk measures of tumor content.

These findings also have potential translational relevance. One persistent challenge in conjunctival melanocytic disease is distinguishing reactive change, CMIL and early invasive melanoma in lesions that may be heterogeneous or borderline by routine histopathology (1). The stage-associated molecular features identified here raise the possibility that spatial biomarkers could complement conventional diagnosis by identifying whether a lesion is dominated by inflammatory activation, transitional remodeling or a melanoma-supportive niche. In that sense, the value of these markers may lie less in identifying single genes in isolation than in defining a spatial molecular context for stage assignment and risk assessment. Such an approach could eventually improve classification, surveillance and perhaps identification of lesions at greatest risk of progression (34–36).

This study also has broader methodological significance. For rare tumors such as CM, where cohort size is limited and tissue is scarce, the ability to extract biologically coherent information from targeted spatial assays is particularly valuable. Our results suggest that with appropriate segmentation and region-aware analysis, even modest-complexity datasets can yield mechanistic insight into disease organization. That principle may be relevant not only to CM, but also to other rare ocular and mucosal tumors for which large-scale transcriptomic resources are unlikely to become available quickly (36, 37).

Several limitations remain. The cohort is small, the panel is targeted, and the analysis is cross-sectional. The framework proposed here should therefore be viewed as a model of stage-dependent tissue states rather than direct evidence of temporal progression. Larger cohorts, broader transcriptomic coverage and correlation with clinical outcome will be needed to determine whether the spatial program identified here can robustly predict progression risk or treatment response (38). Even so, the present work establishes a first-in-field resource and provides a foundation for molecular staging of conjunctival melanoma. More generally, it suggests that the pathobiology of this disease may be best understood through its spatially organized interactions between malignant cells and their local niche.

## Statements

## Acknowledgement

Support for this work was provided by the unrestricted Research to Prevent Blindness (RPB) departmental fund to Scheie Eye Institute, Penn Skin Biology and Diseases Resource-based Center funded by NIH/NIAMS grant [P30-AR069589]. The funder had no role in the design, data collection, data analysis, and reporting of this study.

Support for this work was provided by the Center for Molecular Studies in Digestive, and Liver Diseases [P30DK050306, RRID: SCR_022420] at the University of Pennsylvania Perelman School of Medicine. The funder had no role in the design, data collection, data analysis, and reporting of this study.

Computation resources were generously provided by the Penn Medicine Academic Computing Services (PMACS), whose High-Performance Computing (HPC) and Archive systems were funded by NIH grant 1S10OD012312 NIH.

## Statement of Ethics

Please address the following aspects in your Statement of Ethics.

<u>Study approval statement</u>: The study has been granted an exemption, category 4, by the Institutional Review Board at the University of Pennsylvania (854013).

<u>Consent to participate statement</u>: This study has been granted an exemption from requiring written informed consent by the Institutional Review Board at the University of Pennsylvania (854013).

## Author Contributions

YSH, TM & VL performed sample collection and processing. JM and YC lead data processing and analysis. JM, BD, AC and MR performed the data processing and analysis. BD & MR performed the Cellpose segmentation analysis. JM, YSH, YC & VL drafted the manuscript.

## Data Availability Statement

The raw data set of CM biopsy samples generated by the Xenium Analyzer and analyzed during the current study is available on Zenodo with DOI https://doi.org/10.5281/zenodo.20813650.

## Code Availability Statement

The code to reproduce the results of this paper is available at this Github repository: https://github.com/yuyanchenglab/manuscript_998_prj2401.

