## Supplementary figures and images for "Spatial Transcriptomics Recontextualizes the Cellular Environment of Conjunctival Melanoma"

### Supplemental Figure 1

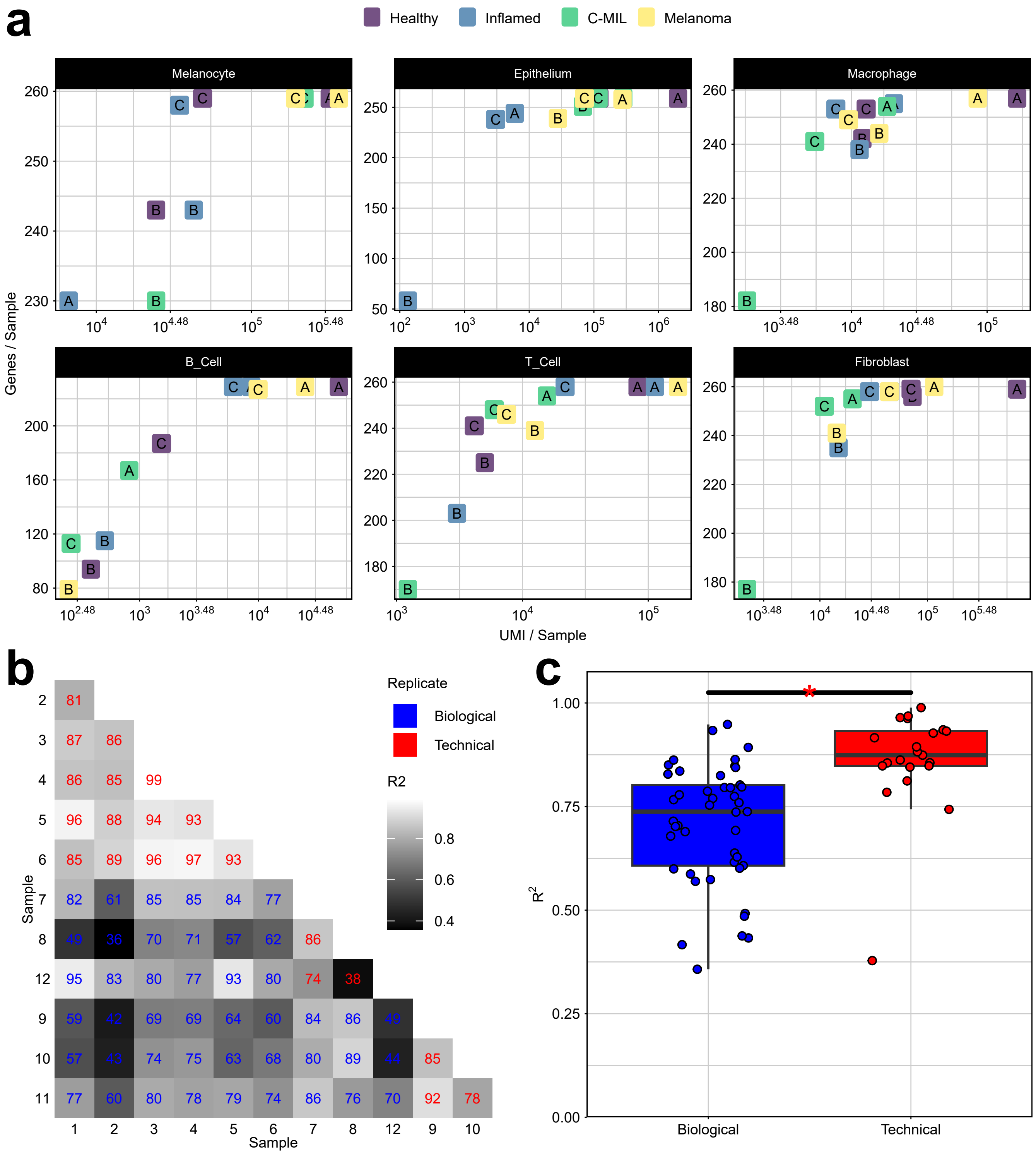

### Supplemental Figure 2

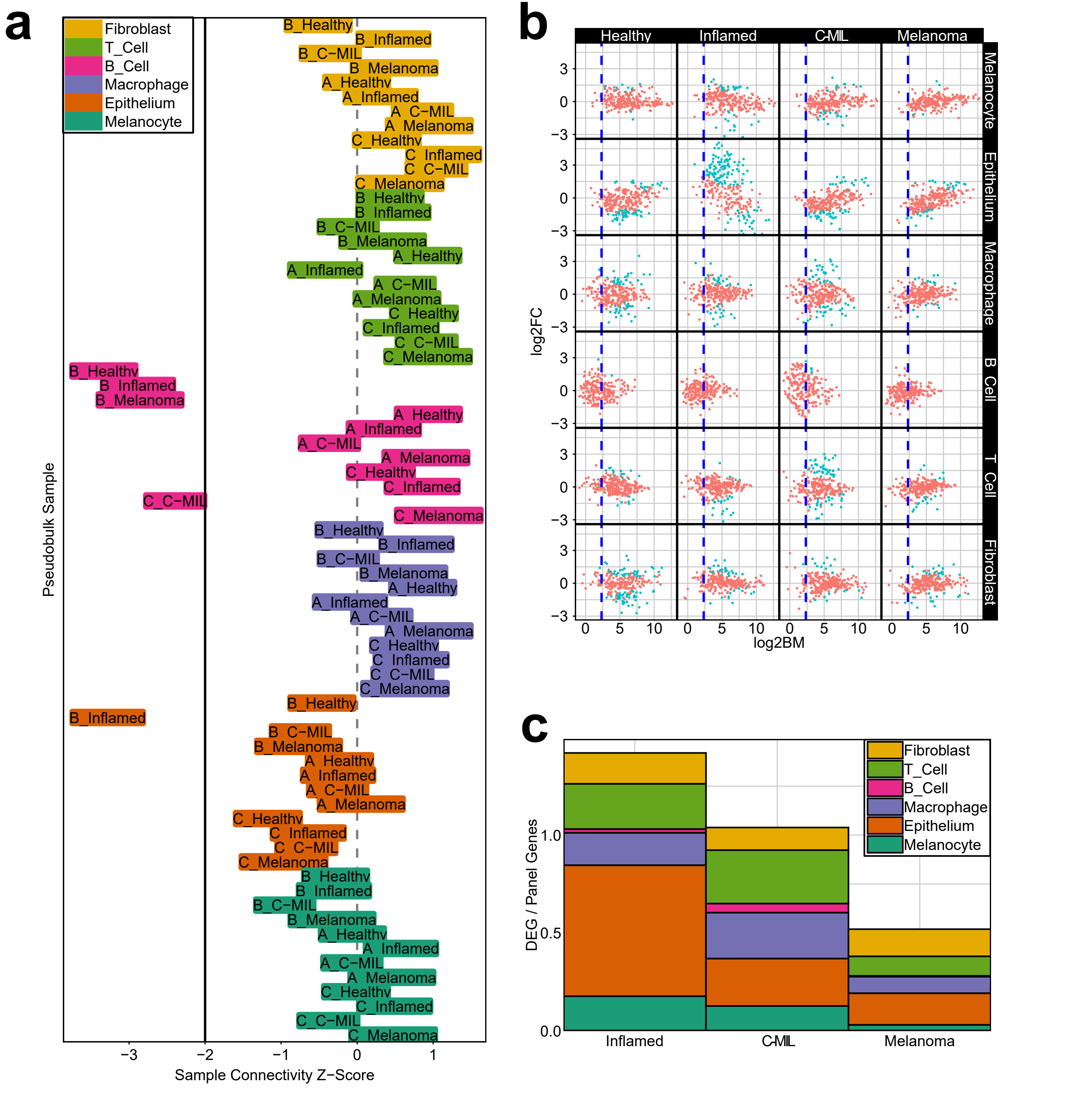
